# Introducing the 4Ps Model of Transitioning to Distance Learning: a convergent mixed methods study conducted during the COVID-19 pandemic

**DOI:** 10.1101/2021.03.23.21254165

**Authors:** Farah Otaki, Shroque Zaher, Stefan Du Plessis, Ritu Lakhtakia, Nabil Zary, Ibrahim Inuwa

## Abstract

Significant concern has been raised regarding the effect of COVID-19 on medical education. The aim of this study was to shed light on the distance learning experiences of medical students and their instructors. A convergent mixed methods approach was utilized. Qualitative and quantitative data was collected using a survey.

The percentage of the total average of satisfaction among stakeholders was 76.4%. The qualitative analysis revealed several themes. This study introduced the 4Ps Model of Transitioning to Distance Learning. It would be useful to leverage the lessons-learned to tailor blended medical programs, with a reasonable mélange of experiences. The study also contributes to the mixed methods research through showcasing a means of adapting it to evaluate critical situations reliably and rapidly.

## Introduction

The novel Corona Virus Disease (COVID-19) pandemic has forced universities around the world to take immediate action to move to delivering courses via online platforms. High quality distance learning typically takes months, if not years, to get off the ground, but COVID-19 forced institutions to make that transition in a matter of weeks (1). Institutions that had already embraced online education, and digitized learning and teaching, were at an advantage at the onset of the pandemic. Having a robust instruction design team, working closely with the academic staff, proved to be key to effectively transitioning to distance learning (2).

Distance learning is proven to be an effective method of acquiring knowledge (3), and provides opportunities for students to work independently, expand their agency, and learn to use novel tools and strategies. It does, however, solicit concerns around matters such as: student engagement in a virtual environment, as well as participation in discussions, where the transition from the workplace or medical school setting to home could result in isolation, and in struggles with establishing boundaries between work and home, which could affect students, faculty, and support staff (4).

Some courses, by their very nature (e.g., those aimed at developing clinical skills), are more difficult to be adapted to an online format, resulting in understandably high levels of stress and uncertainty for both students and instructors. Accordingly, concerns were raised regarding the effect of COVID-19 on medical education, especially in relation to the medical students who have been in the process of preparing for or undertaking assessments that require clinical exposure (5). Also, many students are transitioning from preclinical to clinical stages which is already associated with high levels of student anxiety and uncertainty (6).

However, adoption of online education during the pandemic has shown that it is possible to achieve a number of teaching objectives virtually, particularly for pre-clinical students who have had their entire curriculum moved to online formats (7). The pandemic introduced novel methods of delivering education to medical students (e.g., online webinars via zoom and virtual dissection sessions). Such advanced technological approaches have the potential to maximize engagement among medical students (8).

There are many helpful documents in the literature aimed at supporting institutions undergoing such abrupt transitions, focusing on how to design learning environments, pedagogies, and strategies to engage learners (1, 2, 9-11). One reference emphasized the importance of not comparing emergency remote instruction to established online learning under these circumstances (2). The importance of organizational structures in facilitating critical collaborations between students, faculty, and digital information departments during such turning points are emphasized in the literature (12).

At the College of Medicine (CoM) at the Mohammad Bin Rashid University of Medicine and Health Sciences (MBRU), the transition to online learning took place in March 2020. As is the case with many universities, MBRU is focused on continuously improving andragogical strategies. It is also interested in developing and maintaining Instructional Design (ID), which is defined as a system of procedures for developing education and training programs in a consistent and reliable fashion (13). ID models such as that of ‘Morrison-Ross-Kemp’ (14, 15) consider instruction from the perspective of the learners. Whilst acknowledging the challenges posed by the rapid transition to distance learning, the emergency instructions were still framed as much as possible in alignment with these preset guidelines.

From this perspective, the aim of this study is to reflect on the distance learning experiences of the undergraduate medical students and their instructors at MBRU during these unprecedented times, which is expected to provide valuable insights that can inform future learning and teaching. Accordingly, in this study, we strive to address the following research questions: how was the rapid transition to distance learning, due to COVID-19, perceived by undergraduate medical students and instructors, and how do those perceptions relate to one another? How can we leverage the lessons learned from this experience for MBRU and for other similar higher education institutions?

## Materials and methods

### Context of the study

On 8^th^ March 2020, all educational activities in the UAE were suspended temporarily due to the onset pf COVID-19. The CoM at MBRU transitioned all educational activities (Teaching, Assessment, and Administration) to the online environment, resuming activities after two weeks (as of the 22^nd^ of March 2020), with all employees (faculty and staff) working remotely. The rapid transition was regularly punctuated by policy guidance within the country’s health and education regulatory framework and involved four interrelated aspects. The first one involved supporting faculty members in delivering the content, which involved raising their awareness of available resources that are worth leveraging and offering them a series of relevant learning and development opportunities. The second aspect involved managing the curriculum changes. The transition also involved several measures to facilitate the students distance learning experiences, which included assuring their technical readiness for the transition, including but not limited to the quality of their internet connectivity, maintaining connectedness through instilling open communication channels and continuous engagement, and protecting their health and wellbeing. The last aspect of the transition related to transitioning all assessments to the online environment (16).

The Bachelor of Medicine, Bachelor of Surgery (MBBS) program of CoM is a six-year undergraduate program following a spiral curriculum and divided into three phases. The first academic year constitutes Phase 1 and introduces students to basic concepts in medicine. The second and third academic years represent Phase 2 where teaching is organized around body organ-systems and integrated with clinical medicine. The fourth through sixth academic years constitute Phase 3, through which the students undergo their clinical rotations followed by an internship as a wrap-up.

The transition took place 8 weeks into the 15-week second semester of the medical program. Phases 1 and 2 students had just completed their mid-term In-Course Assessments (ICA) with the year 4 students (i.e., the only cohort in Phase 3) midway through their 4^th^ of a total of 5 clinical rotations (16-18).

### Research Design

Ethical approval for the study was granted by the MBRU, Institutional Review Board (Reference # MBRU-IRB-2020-032). A convergent mixed methods study design was utilized to develop a systemic understanding of the stakeholders’ perceptions regarding the rapid transition to distance learning. Quantitative and qualitative data was concurrently collected and analyzed. The integration of data sources (i.e., students and instructors) and types (i.e., quantitative and qualitative) is meant to raise the validity of the generated findings and relied on joint model analysis (19-21).

### Data Collection

The data was collected using a contextualized version of a validated survey (22, 23). The version of the survey, adapted for this study, aimed at assessing the perception of students and instructors regarding the rapid transition to distance learning due to COVID-19, and its effect on the learning and teaching at the CoM.

The adapted survey was composed of four segments. The first segment is a Likert-type scale of five points (1: Strongly Disagree, 2: Disagree, 3: Neutral, 4: Agree, and 5: Strongly Agree) across 7 components, as per Table 1. Out of those 7 components, 3 were replicated as is for both students and instructors, 2 other students’ components were replaced for the instructors with corresponding alternatives designed to reflect the other side of the same coin, and 2 were unique to the students.

**Table 1.** Description of the first segment of the survey

| Variable | Students | Instructors |
| --- | --- | --- |
| 1 | The transition to the online environment was clearly explained. |  |
| 2 | The technology used in the online environment worked effectively. |  |
| 3 | Adequate opportunities to express my viewpoints and questions were offered to me, during the distance learning. | The University provided me with adequate and timely support throughout the distance teaching. |
| 4 | The online courses' materials were easy to access. | The courses' content and materials were easy to share online. |
| 5 | The online courses' materials suitably contributed to my learning. | - |
| 6 | The online courses' materials available were adequate to meet my learning goals. | - |
| 7 | Overall, I was satisfied with the distance learning. |  |

Legend: This table shows the similarities and differences between the two surveys that were disseminated to capture the perceptions of the students and instructors, respectively. Components 1, 2, and 7 were common for both surveys. Components 3 and 4 were meant to constitute two sides of the same coin. As for components 5 and 6, they were unique to the students.

The second section targeted two out of the four participating cohorts, namely: Year 3 and Year 4. The question asks the respective students to rate, on a scale of zero to ten, the extent to which the distance learning prepared them for the upcoming clinical clerkships. For the students in Year 4, it would be their second clinical sciences’ year. As for the Year 3 cohort, they are transitioning from Phase II to Phase III (hence, from basic to clinical sciences).

The third section of the adapted survey entailed the following two dichotomous questions (Yes/ No), each followed by a separate open-ended question giving the participant the choice to elaborate:

- The transition to the online environment, in response to the COVID-19, significantly impacted my learning (or my teaching) in these courses.
- The transition to the online environment, in response to the COVID-19, significantly impacted the courses’ structure and delivery.

As for the last section of the survey, it was designed to be exploratory to solicit for qualitative data using the following open-ended questions:

- What were some of the advantages of transitioning to distance learning?
- What were some of the challenges that you faced due to transitioning to distance learning?
- Please reflect upon aspects of the alternative modes of instruction deployed that were particularly supportive of your learning (for the students’ distance learning) during the COVID-19 pandemic.
- What aspects of those alternative modes of instruction would you like to sustain in the long run (even after reverting to regular face-to-face sessions)?

In this data collection initiative, no personal identifiers were recorded. Participation was completely voluntary. The privacy of the students and the data confidentiality were protected. The survey was assembled throughout June 2020. In the respective academic year, CoM had 115 in-house and adjunct instructors, and was serving a total of 197 students, across four cohorts Classes of 2022, 2023, 2024, and 2025 (i.e., Years 4, 3, 2, and 1, respectively). Out of those 197 students, as per Table 2, 83 responded (i.e., overall response rate= 42.13%, with the response rates varying across cohorts).

**Table 2.** Response rates across cohorts

| Cohort | Number of Responses | Total Number of | Response |
| --- | --- | --- | --- |

|  |  | Students | Percentage |
| --- | --- | --- | --- |
| Year 4 | 20 | 47 | 42.55% |
| Year 3 | 19 | 35 | 54.29% |
| Year 2 | 16 | 55 | 29.09% |
| Year 1 | 27 | 60 | 45% |
| Total | 83 | 197 | 42.13% |

As for the instructors, a total of 39 faculty members responded (i.e., response rate= 33.91%). Each of the 122 participants were given a unique identification number. The unique identification numbers were complimented with ‘S’ for the 83 students, and ‘I’ for the 39 instructors (i.e., participants 1 through 83 are followed by ‘S’, and 84 through 122 by ‘I’).

## Data Analysis

### Quantitative Analyses

The quantitative data was analyzed using SPSS for Windows version 27.0. The descriptive analysis constituted of computing an overall score of satisfaction for both stakeholders combined (i.e., across the 5 components that are common to both stakeholders), along with a score of satisfaction for the students (i.e., across all 7 components) and another one for the instructors (i.e., across the 5 components that constituted the instructors’ tool). Then, the mean and standard deviation for each of the components of the tools, and the scores: combined, and students and instructors, were then calculated. The validity tests of Cronbach’s Alpha and the Principal Component Analysis (PCA) were performed to ensure the internal consistency and check external variance, respectively, of the adapted tool.

To select the appropriate comparative analyses tests, a test of normality was conducted for each of the 7 components, and for all three scores of satisfaction (combined, and students and instructors). The data of each of the 7 components, independently, and the combined score of satisfaction, and that of the students and instructors, all turned out to be not normally distributed.

Accordingly, the Mann-Whitney test was used to compare the combined score of satisfaction, and each of the 5 common components independently, between both groups of stakeholders (students and instructors), and the combined score of satisfaction, and that of the students and instructors, between those who answered ‘Yes’ (versus those who answered ‘No’) to each of the two dichotomous questions of the third section of the survey. The potentiality of association between the perceived readiness for transition for students of Classes of 2022 and of 2023 (i.e., Years 4 and 3, respectively) and the dichotomous variables was also assessed using the same test. In addition, Chi-squared was used to assess any potential associations between the two dichotomous variables of the third section of the survey, and the group of stakeholders and the cohort of students.

Finally, Bivariate Spearman Correlations were conducted to assess the extent to which the combined score of satisfaction, and that of the students and instructors, can be explained by changes in the stakeholders’ perception of the components of the scores, and weather the perceived readiness for transition for students of Classes 2022 and 2023 (i.e., Years 4 and 3, respectively) is associated with the students’ score of satisfaction and/ or the components of the respective score.

### Qualitative Analysis

The data collection phase was completed before the start of the data analysis. Thematic analysis by five researchers (SZ, SDP, RL, NZ, and II) was carried out. The factors that could influence the perceptions of the researchers, in relation to the subject matter, were recognized upfront. The qualitative data was divided into five datasets: one for each of the four cohorts of students and one encapsulating the data of all the participating instructors. The research process was inductive, based on the constructivist epistemology. The consistency, in relation to the theoretical assumptions, was assured throughout the study, where one member of the research team (FO) facilitated, and controlled for the uniformity and steadiness of the qualitative analysis process, without engaging in the actual inductive analysis. This interpretative approach enabled the researchers to gain a thorough understanding of the phenomenon under investigation (i.e., rapid transition to distance learning at CoM at MBRU).

The six-step framework initially introduced by Braun and Clarke (2006) was adapted (24). This multi-staged approach to thematic analysis has been used repetitively in research concerning health professions education (25, 26). NVivo software version 12 plus (QSR International Pty Ltd, Vic, Australia) was used to assign codes to the text fragments, and in turn expedite the classification of the data into categories and themes.

The analysis process started with the researchers acquainting themselves with the data, where they collectively skimmed through all the datasets and reflected upon them. Then, as the second step of the adapted approach, the text fragments that refer to the same aspect of the distance learning experience were compiled together, labelling each with an all-encapsulating title. This was done for each of the five datasets separately (each researcher was randomly assigned one of the five datasets). This is how the qualitative data was examined line-by-line, while the researchers were assigning codes to text fragments, until data saturation was attained.

The resulting categorization schemes for the five datasets were mapped onto each other to compare perceptions. The researchers reflected upon areas of harmony and discord within and across the datasets.

Following that, the discrete concepts that surfaced from the independent, concurrent analyses underwent several rounds of reflections. The multiple ways by which the concepts could relate to one another were identified. This led to the generation of categories that extensively cover all that surfaced in relation to the research questions, which set the stage for the researchers to work on step three of the adapted thematic analysis approach. The researchers examined the categories, again, to find the best way to merge them into higher order themes.

For the fourth stage, the generated themes and categories were then reviewed to ensure that the data within each grouping are sufficiently similar, and data in between the clusters are distinct enough to deserve isolation. To complete stage, the researchers agreed on labels and documented explanations for all the themes and categories. This constituted the basis of the study’s conceptual framework which guided the last step of reporting upon the findings (27).

### Joint Model Analysis

The findings from both types of analyses: quantitative and qualitative, were merged to result in a meta matrix. This mixed methods integration was done using joint display analysis which ultimately led to meta inferences (28, 29). The findings of both analyses were compared (and contrasted) to weave together a meaningful narrative. The researchers looked for areas where the findings of the two analyses confirmed each other. They also examined areas that were unique to one analysis (quantitative or qualitative), with the intention of building upon the generated insights, and investigated whether these unique areas complement the areas that were confirmed by both analyses. Throughout the iterative integration process, the researchers were open to potential discordances between the findings of the two analyses.

## Results

### Quantitative Analyses

The reliability score of Cronbach’s Alpha for the tailor-made evaluation tool, that captured the perception of the stakeholders was 81.8%. The percentage of the total average of the students, instructors, and both groups of stakeholders combined were 73%, 81.64%, and 76.4%, respectively, as per Table 3.

**Table 3.** Output of descriptive quantitative analysis

| Stakeholder: | Students<br>(7 Components) |  |  | Instructors<br>(5 Components) |  |  | Combined<br>(5 Components) |  |  |
| --- | --- | --- | --- | --- | --- | --- | --- | --- | --- |
| Identification Number<br>of Item of Satisfaction | Mean (SD) | Percentage<br>of the<br>Mean | Category | Mean (SD) | Percentage<br>of the<br>Mean | Category | Mean (SD) | Percentage<br>of the<br>Mean | Category |
| 1 | 3.39(1.16) | 67.80% | N-A | 3.90(0.93) | 78.00% | A | 3.56(1.11) | 71.20% | A |
| 2 | 3.96(0.91) | 79.20% | A | 4.18(0.84) | 83.60% | A-SA | 4.03(0.89) | 80.60% | A |
| 3 | 3.83(1.11) | 76.60% | A | 4.18(0.93) | 83.60% | A-SA | 3.94(1.06) | 78.80% | A |
| 4 | 3.94(0.99) | 78.80% | A | 4.10(0.87) | 82.00% | A-SA | 3.99(0.95) | 79.80% | A |
| 5 | 3.56(1.15) | 71.20% | A | - | - | - | - | - | - |
| 6 | 3.54(1.12) | 70.80% | A | - | - | - | - | - | - |
| 7 | 3.35(1.08) | 67.00% | N-A | 4.05(0.82) | 81.00% | A | 3.58(1.05) | 71.60% | A |
| Score of Satisfaction (7<br>components): | 25.57(5.91) | 73.06% | A | - | - | - | - | - | - |
| Score of Satisfaction (5<br>components): | 18.48(4.15) | 73.92% | A | 20.40(3.54) | 81.64% | A-SA | 19.11(4.05) | 76.40% | A |

N=Neutral, A= Agree, and SA= Strongly Agree

According to the PCA (Kaiser-Meyer-Olkin Measure of Sampling Adequacy), 73.6% of the variance can be explained by the instrument which means the instrument is not only reliable but also, according to Bartlett’s Test of Sphericity, valid to measure what it is intended to measure (p<0.001). Along the same lines, the Bivariate Spearman Correlations showed how the changes in each of the three scores can be explained by changes in all the respective components.

The instructors, with a mean of satisfaction of 20.40(3.54), rated the distance learning experience at CoM higher than the students, with a mean of satisfaction of 18.48(4.15) (p= 0.015). In addition, the instructors were significantly more satisfied than the students in relation to the following two components: “the transition to the online environment was clearly explained” and “overall, I was satisfied with the distance learning” (p= 0.022 and 0.001, respectively). As for the remaining three components, common between the two groups of stakeholders, there was no significant differences. In addition, there was no significant difference between the two groups of stakeholders in relation to their perception of whether, or not, the transition significantly impacted the courses’ structure and delivery. Yet, the students perceived the transition to significantly affect their learning more than the instructors perceived the transition to affect their teaching (p= 0.002).

There seemed to be a statistically significant difference between the score of satisfaction between the cohorts within the students’ group of stakeholders (p= 0.001), with Year 1 having the lowest mean of satisfaction of 22.67(6.27), followed by Year 3 with a mean of satisfaction of 24.26(5.22), then Year 4 with mean of satisfaction of 27.40(4.59), and finally Year 2 with a mean of satisfaction of 29.75(4.52). As illustrated in Figure 1, there was also statistical significance between the four cohorts across the 7 components of the tool (p<0.05). Moreover, across the cohorts, there was no significant difference between the students’ perception of whether, or not, the transition affected their learning in the courses. Yet, there turned out to be statistical significance across the cohorts in relation to how the students perceived the transition to have affected the courses’ structure and delivery (p= 0.018).

**Figure 1.**
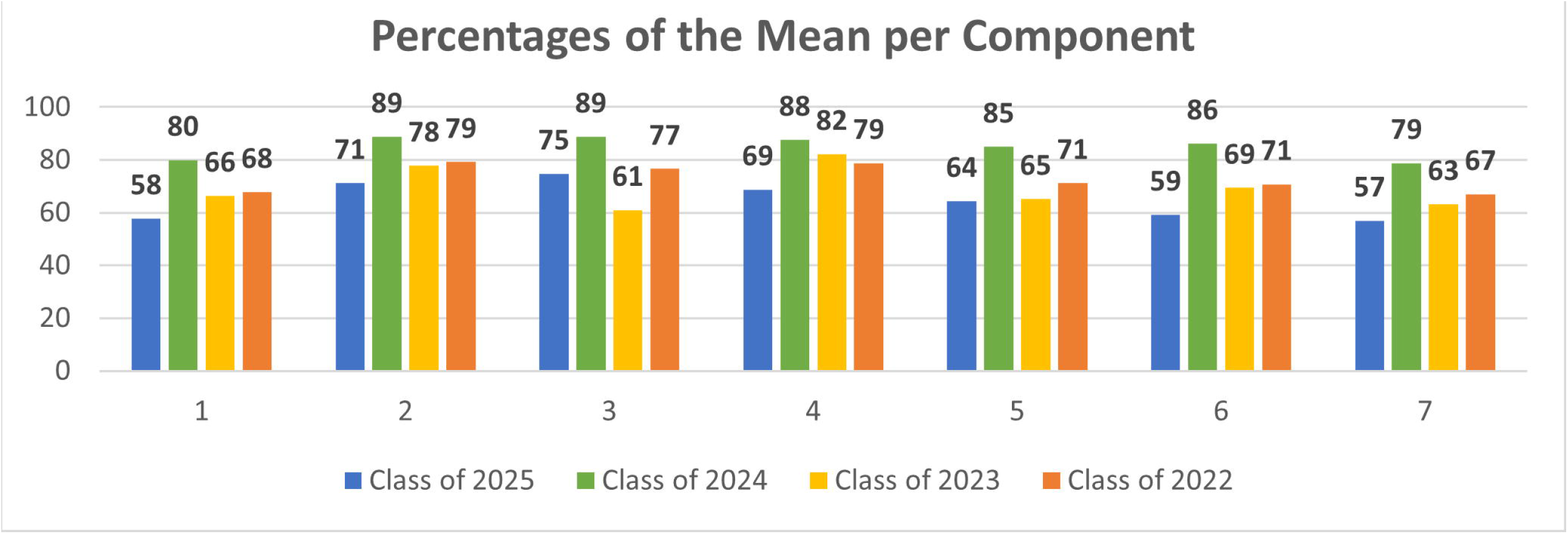
Percentages of the mean for each component across the cohorts of students (Class of 2025= Year 1, Class of 2024= Year 2, Class of 2023= Year 3, & Class of 2022= Year 4).

In relation to the level of perceived readiness to transition to the clinical years (i.e., Year 4 to Year 5, and Year 3 to Year 4), students in both cohorts rated themselves low (on the zero to ten scale) with a mean of 3.97(2.32), with students of Year 4 perceiving themselves to be significantly more ready than those of Year 3 (p= 0.004). The Bivariate Spearman Correlations showed that the level of perceived readiness is associated with the students’ score of satisfaction, and with 6 out of 7 of the components of the evaluation tool (p<0.05). The only component which turned out not to be associated with the perceived level of readiness is “the online courses’ materials available were adequate to meet my learning goals”. The level of perceived readiness was also not associated with whether, or not, the students observed a change in the learning, and courses’ structure and delivery.

The analysis also showed that there is an association between the scores of satisfaction (students and instructors, and combined) and whether, or not, the respective groups of stakeholders perceive the transition to have impacted the learning or teaching (p= 0.012) and the courses’ structure and delivery (p= 0.003), where the stakeholders who were more satisfied were significantly less likely to notice a change in the learning or teaching, and the courses’ structure and delivery.

## Qualitative Data

The qualitative analysis of the perception of the students and instructors showed an interplay between two interlinked themes: People and Processes. The stakeholders perceived this interaction to take place on the Platform, which constituted the third theme of the analysis, all of which is guided by the last theme, namely: Policies. These four themes came together as per Figure 2 to constitute the study’s conceptual framework: 4Ps Model of Transitioning to Distance Learning.

**Figure 2.**
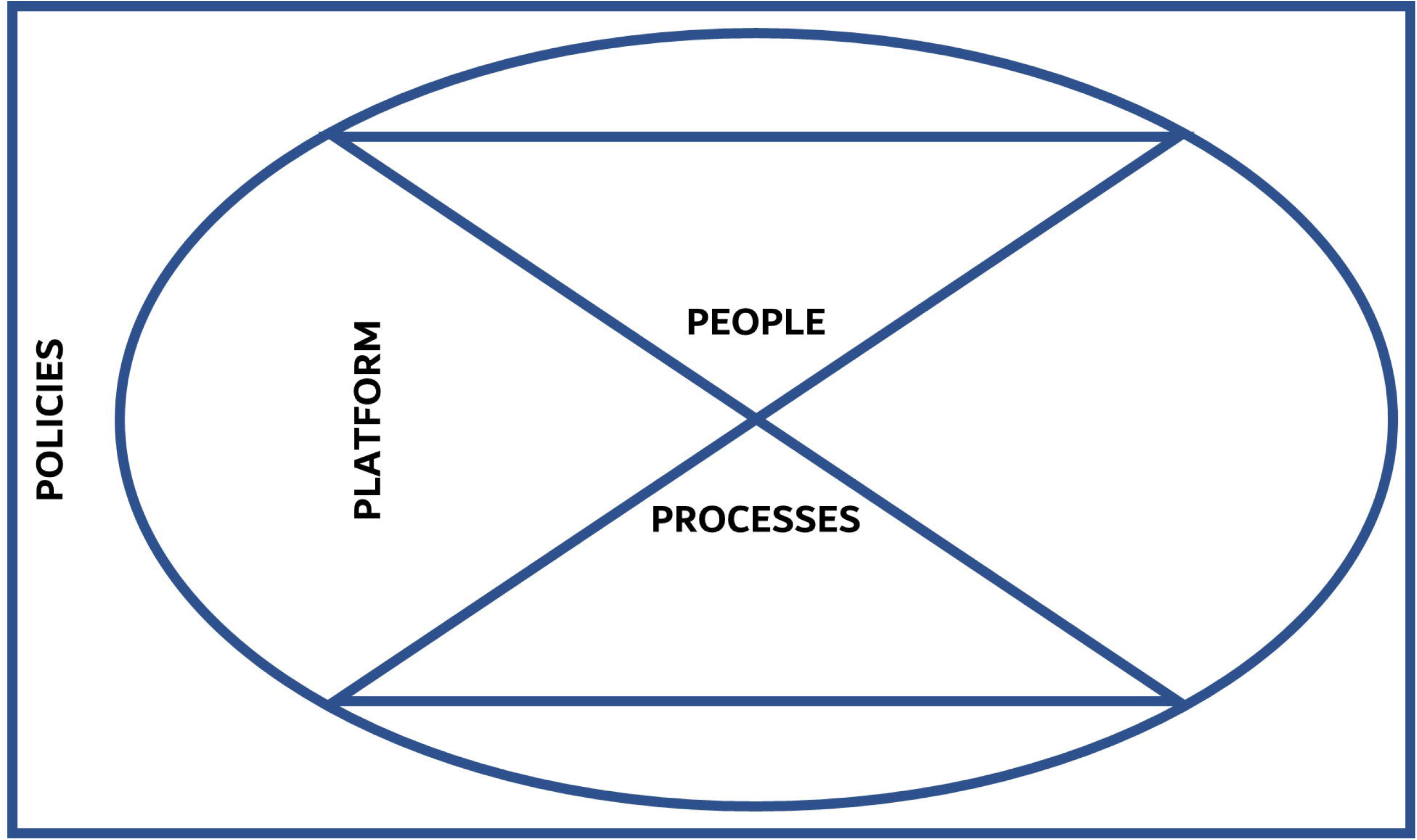
The study’s conceptual framework: The 4Ps Model of Transitioning to Distance Learning

### Theme 1: People

This theme refers to text fragments that pinpoint variables on individual or interpersonal levels that seemingly affected the perceived quality of the distance learning experience, the value obtained from the experience, and/or the stakeholders’ level of satisfaction with the transition.

On an individual level, the transition to distance learning affected the stakeholders differently. Some students highlighted that it increased the level of stress:

> 25-S-Y1: “…I felt more overwhelmed and stressed in comparison to how I felt when we used to have classes at the University. I was required to learn everything on my own, and that is not an easy task…it became more difficult to approach the professors in comparison to when I used to be on campus…”
>
> 61-S-Y4: “…days became monotonous, and with the lockdown and minimal engagement with the outside world, my motivation and discipline got affected, even my mental health…”

The students, especially those of Year 3, were particularly concerned about the absence of experiential clinical training:

> 52-S-Y3: “…learning about physical examination and history taking online, as opposed to actually doing them, was insufficient; I do not feel competent to do them in a clinical setting…”

Similarly, some instructors highlighted challenges that they faced due to the transition.

> 120-I: “…the working hours extended to overlap with the resting hours… I noticed some students experienced online classroom fatigue… at some point, we lost the boundaries between work and home; work and rest somehow overlapped…”

Some students expressed that the transition offered them a set of advantages and lessened the pressure on them:

> 55-S-Y1: “…I honestly felt like this has been a much-needed break. I do not feel peer-pressured (for lack of a better term). I am much more relaxed now, as I have struggled with this, prior switching to remote learning, given that most of the students around me are high achievers…”
>
> 11-S-Y4: “…I think I am performing better as distance learning allowed me to have more time on my hands. I organized my timings better than when I was experiencing ‘regular’ university days. I enjoyed a nice balance between studying, family time, and hobbies…”

It was also clear to the instructors that the transition to distance learning was an opportunity to empower the students, encouraging them to be proactive and to engage in active learning.

> 117-I: “…it allowed for active, collaborative learning… the students identified their own learning needs…we discovered that via the online platform we can more effectively engage the students with the courses’ content…’’

The instructors also indicated other advantages, most of which were on an individual level.

> 93-I: “…we benefitted a lot. We learned new teaching modalities. The transition certainly led to greater engagement as some of the quieter students felt more confident to ask their questions using the chat box…”

The instructors perceived the transition to have hampered the interactional aspect of the learning process. To some instructors, the relationships that they have with the students constitute the highlight of their job.

> 88-I: “…in the online environment, there is a ‘disconnect’ between student-teacher interactions and it is difficult to ensure the students stay involved…”

A lot of the students, especially the more junior ones, also yearned for more interaction.

> 35-S-Y1: “…the discussions that I used to have with my fellow classmates, during the sessions, were very beneficial; they constituted opportunities to learn from one another and clear-up any misconceptions about the lecture…”

Some students especially the more senior cohorts, mainly those of Year 4 but also those of Year 3, saw interactional value in the transitioning.

> 68-S-Y3: “…we did not have to get up too early or go back too late which made learning easier for us and more joyful since we got the chance to spend quality times with our families…”

### Theme 2: Processes

This theme included the text fragments that refer to the core of the system of distance learning and encapsulated three categories, namely: learning and teaching, assessment, and organization and delivery.

A lot of students perceived the transition to be smooth, and for the didactic learning and teaching, and assessment to be of the same quality, or in some cases even better, relative to that in a regular face-to-face set-up.

> 25-S-Y1: “…there were a variety of teaching methods that were put out for all students to benefit from…I valued having pre-recorded lectures, and virtual discussions on topics accompanied with question-and-answer sessions and regular feedback sessions…”
>
> 24-S-Y2: “…the schedule was flexible, and I was able to go through the lectures at my own pace. I was able to take detailed notes and repeat the points that I have missed…”

The lack of experiential clinical training left a prominent gap, from the perspectives of the entire student body and that of the instructors.

> 7-S-Y2: “…the Foundations of Clinical Medicine were affected the most. The course was adjusted, of course…it still is not the same as seeing a Simulated Patient…”
>
> 85-I: “…nothing can replace clinical skills training with real patients which is the core of medical teaching…simulation and clinical-based teaching could not be performed effectively. There were no interactions with patients…”

The students of Year 4, relative to the rest of the students, were more grounded, and at ease with the absence of experiential learning and had faith that they would get opportunities to make-up for it in the future.

> 20-S-Y4: “…the only missing component is clinical exposure, which we will get plenty of opportunities to make up for in the upcoming years…”

Some students highlighted that they needed to adapt their learning styles to cope with the changes.

> 34-S-Y1: “…I had to figure out a whole new way of studying which is something I had already done at the beginning of this semester to become more accustomed to learning anatomy. I had to do it all over again because I needed to find a new way of studying that is more appropriate to distance learning, and that took some while…”

A few students believed that the transition impeded, for differing reasons, their performance in assessments.

> 68-S-Y3: “…I think the exams were affected negatively because we were barely given any input as to what to expect, which in some cases was frustrating. As a student, I need to know what lectures I will be tested on, or at least how many questions per course, all of which was transparently shared with us previously…”

Some students highlighted the impact that this had on the quality of feedback provided to them following exams.

> 80-S-Y3: “… in this course, we were not given adequate feedback neither for in-course nor for the final assessment…”

Others felt that doing the assessments from the comfort of their homes was more convenient.

> 33-S-Y2: “…during the assessments that we underwent online, I noticed I was much calmer. I am usually very stressed in the exam hall, and so from this perspective, the transition constituted a big advantage…”

Students, especially those more junior, faced plenty of challenges around the multiplicity of platforms and the scheduling application.

> 23-S-Y2: “…the multiplicity of channels and approaches was a bit confusing. Some courses were prerecorded, and others were real-time on teams or zoom. At some instances, it became very hard to keep-up with all that was going on online…”

Others felt that the workload increased due to the transition.

> 16-S-Y4: “…the quantity of the learning material significantly increased in an attempt to compensate for missed clinical hours…”

There were some stakeholders who highlighted the advantages of the transition in terms of program organization and delivery.

> 50-S-Y3: “…the learning process has been much more efficient in didactic courses than in university. The professors were all very receptive to feedback and questions…”
>
> 110-I: “…distance learning compelled me to try other modes of delivery. Distance learning also proved that not all lectures need to be delivered in the college within the confines of the classroom. We now have a repository of online material that can be accessed by students…”

### Theme 3: Platform

The students and instructors repeatedly referred to the medium through which the distance learning occurred. They highlighted how the tangible and the intangible came together to allow for the experiences to occur. Within this theme, two categories were grouped: the virtual environment and Information Technology (IT).

The transition to the virtual environment has been a challenge to a lot of the students, especially those of Year 1. This group of students had just started their higher education path when the outbreak started, which exacerbated the perceived uncertainty.

> 34-S-Y1: “…this was a completely new territory for me. At the beginning, it was strange to learn in this environment. The fact that we had to learn the hardest two courses of the semester and to build our foundation, in these new circumstances, was quite tough…”

Despite the perceived uncertainty that all stakeholders were experiencing, the students expressed appreciation of the virtual environments and online platforms.

> 39-S-Y2: “…they provided a good experience that is as close to the classroom as possible…video-recorded sessions were fantastic.’’
>
> 51-S-Y4: “…in Family Medicine, we had students preparing presentations on topics that they were interested in which helped us maximize the entailed learning. The Team-Based Learning was great, and the effective use of Learning Activity Management System (LAMS) software helped in maintaining the momentum…”

Instructors also highlighted aspects of the online environment that they appreciated.

> 103-I: “…distance learning was a good method for engaging Clinical Adjunct Faculty. The schedules of those stakeholders are extremely packed; it is great to enable them to teach without needing to come on campus… This needs to be maintained on the long-run…”
>
> 106-I: “…it saved time…working from home rather than having to rush to a lecture hall…”

Some students reflected upon the limitations and challenges associated with the utilized platforms and the technical glitches that they faced along the way.

> 36-S-Y3: “…not knowing ‘where’ classes were held, on which platform…some instructors resorted to calling student names to increase engagement. This, at times, solicited my anxiety…
>
> 65-S-Y4: “…I do not think any Information Technology platform will offer a clinical experience that is congruent to what we get from face-to-face interactions and real-life experience…”

### Theme 4: Policies

The last theme is related to the ‘non-negotiables’, be it at the level of the institution or beyond. The pandemic constituted a reality-check. Nations had to rapidly respond by instilling directives that imposed restrictions on all sectors, including but not limited to higher education. Within this sector, universities needed to adapt to the constraints by instilling institutional policies that minimized losses and maximized value, in abidance with external restrictions.

The lockdown, quarantine, and the formal obligation to stay at home were the restrictions most highlighted by the stakeholders. Other ministerial directives, that affected the policies and procedures at MBRU, were also brought-up by the stakeholders.

> 45-S-Y3: “…social distancing is important in these times and it has been crucial to stay at home…”
>
> 19-S-Y2: “…another example is when the ministry issued new grading rules (regarding optional Pass/Fail). There was an understandable delay in communicating to the University and in turn to the students. This generated confusion; the students started speculating which solicited panic…”

The restrictions caused challenges, among which was the inability to conduct Objective Structured Clinical Examinations (OSCE).

> 39-S-Y2: “…OSCE was cancelled and the grades were divided among the rest of the assessing activities; we had a written final exam, which was also a considerable shift from the usual structure of the course…”
>
> 92-I: “…the course material and assessment had to be restructured to account for the inability to conduct an OSCE…”

The other challenges were related to shortcomings of the experiences and/ or the medium in which the experiences are taking place, which could have been circumvented by endorsing, and ensuring effective implementation of, appropriate institutional policies.

> 12-S-Y3: “…the duration of the recordings exceeded the allocated 50 minutes duration of the session…”
>
> 15-S-Y3: “…I would have preferred for sessions that are not interactive to be pre-recorded…”

Most stakeholders expressed satisfaction with how the University applied the external directives, and how the support units provided the instructors with platforms for the alternative pedagogies.

> 9-S-Y2: “…the university did a great job at dealing with this crisis…”
>
> 33-S-Y2: “…the timeline got extended; we ended-up having more time to study and prepare for the assessments, which was a great plus…”

## Mixed Methods Integration

Mapping the output of the quantitative analysis onto that of the qualitative analysis revealed a systemic perspective of the situation, illustrated in the study’s side-by-side joint display (21) (Table 4). The convergence of findings enabled the development of the perceptions of students and instructors, of the transitioning to distance learning, and their interrelationships.

**Table 4.** Side-by-side joint display

| Key Quantitative findings → | ← Key Qualitative findings |
| --- | --- |
| • Stakeholders were satisfied | • <u>People</u>: Personal and Interpersonal variables |
| • Instructors were more satisfied than students |  |
| • Students perceived readiness to transition |  |
| to the clinical years was low |  |
| • Among the students, those of Year 2 were most satisfied, followed by those of Year 4 then those of Year 3, and lastly the students of Year 1, who were the least satisfied |  |
| • Transition did not significantly impact the courses' structure and delivery | • <u>Process</u>: Learning and Teaching, Assessment, and Organization and Delivery |
|  | • <u>Platform</u>: Tangible and Intangible aspects |
| - | • <u>Policies</u>: Institutional, National, and International levels |

On its own, the quantitative analysis showed the attitudinal reactions of the two groups of stakeholders, and how they relate to one another. These findings were confirmed and expanded upon by looking into the output of the inductive thematic analysis. Moreover, the qualitative data analysis, on its own, uncovered details about the platform and the policies, which generated a more holistic perspective of what has been taking place. Through the meta-inferences, generated from the joint model analysis, it became apparent what the strengths and the opportunities for improvement around the experience are.

## Discussion

This study provides a timely reflection on a unique disruption in education in the context of the ongoing COVID-19 pandemic, due to its sudden onset, rapid evolution, and global impact. The consequential abrupt cessation of all intramural educational processes, as human activity came to a grinding halt, assumed historic proportions. The associated biological threat imposed an additional stress and required appropriate mitigation measures. Medical professionals are familiar with rapid responses to emergencies, in general, and infectious outbreaks, in specific. However, as medical educators responsible for nurturing and graduating safe doctors, the term: *risk*, takes on entirely different connotations (30). In medical education, the risk was two-fold: first, in the interruption of the educational process, and second, the exposure to a biological threat of medical students learning onsite in healthcare facilities.

This study found a high degree of satisfaction with institutional measures devised for distance learning and teaching, across the surveyed stakeholders. The strongest voice of the student body endorsed the gained flexibility, the access to pre-recorded sessions and blended methods, and the multiplicity of digital learning tools. The stakeholders also appreciated the emergence of increased peer-collaboration and thrived on the chat box interactions during real-time virtual learning sessions. On the assessment front, the students perceived benefits related to having extended preparatory time towards final examinations. Some students found taking exams in the quiet privacy of their homes de-stressing. Others appreciated the feeling of safety of staying at home during the pandemic, which was complemented by the emotional support provided by their families.

The instructors felt a sense of achievement in their enhanced capability on e-learning tools, adopting technology that they would have been otherwise reluctant to use. Along the same lines, innovation, flexibility, and overcoming resistance to alternative technology-enhanced tools have been endorsed in the literature as transformational in medical education (Wayne, 2020). The robust establishment of distance learning at MBRU was particularly beneficial to off-site clinical faculty who could balance service commitments with academic engagements. Enhanced interaction through innovative techniques, like LAMS, allowed mutual tutor-student satisfaction. Virtual microscopy, already well established in the institution, seamlessly blended with remote learning.

However, students’ satisfaction with the transition to learning was lower than the faculty transition to teaching. This could be attributed to the high level of uncertainty perceived by the students due to their pre-existing anxiety around their performance in assessments and progression. Specific qualitative inputs provide multifactorial explanations for the academic struggles reported by students. Early-stage learners grappled with school to college transition, the brand-new curriculum content of medical foundations, lack of exposure to cadavers, and inability to live up to expectations of self-directed learning, clinical reasoning, and interpretative skills. They also did not get the chance to leverage the resources that the university offers as part of the “university life” to strengthen and build their character and resilience (e.g., student clubs and co-curricular activities). Counsellor support was boosted and proved invaluable in providing some degree of relief.

At the other end of the spectrum, deprivation of on-site, first-hand clinical exposure created anxiety in senior students at the critical juncture from pre-clinical to clinical years. Even in pre-COVID times, the preclinical to clinical transition is considered a period of workload and professional socialization stress (6). The requirement for rapid adaptation induced by COVID isolation was bound to aggravate the situation. In contrast to satisfaction with flexibility (expressed by many students), other students reported a struggle with exercising discipline in organizing their study schedules. Students missed class-room interactions and complained of content overload, affecting coping mechanisms. Loneliness, absence of group dynamics, and balancing an intrusive home environment were psychosocial demotivators. It is worth noting that some of this anxiety might be due to the comparison that naturally takes place between the students in such spiral curriculums; all the years are interconnected and build upon each other, which is why the students tend to evaluate the quality of what they are receiving in relation to what others are going through. The effect of such doubt in readiness and the resulting low level of self-efficacy (and perhaps self-esteem) can affect the students’ professional identity, as well.

Revised assessment weightage and schedules that deviated from original plans, and dissatisfaction with quality of feedback were also areas of concern. The achievement of competence in Entrustable Professional Activities form the cornerstone of graduating safe physicians; their learning and subsequent assessment cannot be overemphasized (Rose, 2020). Yet, the tussle between self-preservation and vulnerability of professional learning can be a moral dilemma (30). An insightful analysis suggests a 3-point solution towards assessment, “focusing on outcomes, broadening the assessment toolbox, and improving the Undergraduate Medical Education (UME)-to-Graduate Medical Education (GME) transition” (31).

The lack of association between the perceived readiness to transition, and the availability and accessibility of resources online, is also worth highlighting. It was clear from the quantitative analysis that the extent of perceived readiness is not associated with the students’ satisfaction with the availability and accessibility of the resources. Although the students were quite satisfied with the online environment, they were acutely aware that there is no replacement (equivalent alternative) to real-life clinical experience and face-to-face interactions with patients. Similar views have been expressed by medical students from Southampton in the UK, who worried about the reduced learning exposure to certain specialties, the effect on preparation for skills-based examinations, and their immediate residency and career prospects (5).

Instructors felt supported in ensuring delivery of content in keeping with curricular outcomes. There were specific challenges with lack of on-screen, interactive exchanges. The modification of teaching clinical skills was clearly a common discontent among instructors and students alike. Despite the professional and smooth conduct of an online OSCE (e-OSCE), specific domains of professional skills could not be tested. The need for competency is not that of the student alone, instructors too must be adaptable, and should learn to engage with the students beyond didactic delivery (32).

Information Technology including hardware, software, and the internet connection, upon which the whole experience was anchored, did not pose a significant challenge in the eyes of the stakeholders. Neither students nor instructors expressed any significant degree of dissatisfaction or disruption associated with the platform. This is of particular interest since it was the university’s first experience with remote proctored exams. Clearly, the IT support to teaching and assessment was a yeoman effort. World-wide, medical institutions have highlighted the efficacy of globalized outreach for continuity of education through digital platforms when their international students returned home during the pandemic (33). Taking a step further, there is potential for inter-institutional collaboration in *teaching beyond borders* as we move forwards.

The perceived efficaciousness of distance learning, accompanied with the lessons learned from this experience, call for sustaining components of what has been deployed at CoM in MBRU irrespective of how matters unfold pertaining to COVID-19. At a macro-level, the insights from the experience urge a revisit of pedagogic management of the human resource-infrastructure interface. For the teacher-learner partnership to evolve under the current circumstances, reflection on the experience, and identification of innovative but feasible alternative routes, adds value on the long-run (4). In terms of students, building self-directed learning capability, peer-collaboration, and access to diverse digital resources are clear winners. Instructors must sustain and step-up in digital teaching, embracing technology, spearheaded by faculty development. Virtual reality and Artificial Intelligence platforms are galvanizing the available options. Sustainability of technology is a clear area of focus.

Uncertainty of IT environment at the remote users’ end is an area of concern and needs deliberation to search for risk mitigation. There is also the need to evaluate and maintain the quality of educational delivery and the long-term outcomes of courses and programs (34). Yet, given the clear gap in clinical experiential learning, which constitutes the core of medical education, there is consensus across the board that full online programs would be limiting. Blended programs that entail a reasonable mélange of experiences foster active learning among students. There have been several identified ways of students’ curricular enhancement through inclusion in community education, telemedicine, and even in patientcare with adequate physical and financial protection which will provide upskilling for future physicians (35). A Strengths, Weaknesses, Opportunities, and Threats (SWOT) analysis of the impact on education of the pandemic highlights, among many of the factors discussed in the preceding paragraphs, the opportunity for educational leadership in times of crisis and the challenge of executing it with budgetary constraints (36).

The COVID-19 experience is also a living example to demonstrate to students the health inequities and difficult ethical decisions in a contemporary life or death scenarios; it constituted an invaluable experience that reinforces knowledge of health systems and standards of practice (7). Resilience, grit, and tolerance have transformed from mere teaching points to demonstrable practice (7). The tag: *front-liner*, has become a proud identity, inspirational for the budding physician.

This study is characterized by several limitations that are worth shedding light on. Although the focus on a single program enabled the development of thorough reflections and insights, the generalizability of the generated findings is limited to institutions that are characteristically and contextually like MBRU. It would be worthwhile for future studies to collect data from several programs and run a comparative analysis. The small sample size and low response rate were also limitations. The data collection extended until after the end of course assessments, and hence, the perception of the participating students might have been influenced by their performance in the respective exams and the accompanying emotions. There was also no differentiator between in-house versus adjunct faculty, and those who teach predominantly basic or clinical sciences-it would be interesting for future studies to collect the type of affiliation and engagement of the respective instructors to see if those variables play a role in the educators’ impression of the overall learning and teaching experience. Finally, it is worth exploring the perception of education leaders in relation to the transition because they are key to sustainable transformation.

## Conclusion

This study introduced the 4Ps Model of Transitioning to Distance Learning, which explain how differing variables related to People, Processes, Platform, and Policies come together to enable the distance learning experience. While the response to the pandemic and the rapid transition to distance learning at CoM, during those unprecedented timings, was swift, plenty of challenges were faced, most of which were effectively circumvented. The virtual learning demonstrated efficacy in many aspects. In addition, the involved stakeholders were offered plenty of opportunities to develop themselves and the systems within which they operate. However, the gap resulting from inability to virtually compensate for experiential learning was evident in this study, since this type of training is integral to medical education and professional identity formation. It is necessary for health professionals’ educators to adapt to new tools and engage in dialogue to maintain effective educational missions in preparation to any force majeure, COVID-19 or otherwise.

## Data Availability

All data referred to in the manuscript is available with the corresponding author and can be shared upon request.

## Acknowledgement

The authors would like to thank Dr. Fatemeh Amir Rad of Hamdan Bin Mohammed College of Dental Medicine (HBMCDM) at MBRU for her support in preparing the project proposal.

